# A Scoping Review of Interventions to De-implement Potentially Harmful Nonsteroidal Anti-inflammatory Drugs (NSAIDs) in Healthcare Settings

**DOI:** 10.1101/2023.07.29.23293362

**Authors:** Michelle S. Rockwell, Emma G. Oyese, Eshika Singh, Matthew Vinson, Isaiah Yim, Jamie K. Turner, John W. Epling

**Author notes:** **Address for Correspondence:** Michelle Rockwell, PhD, RD, Department of Family & Community Medicine, Virginia Tech Carilion School of Medicine, 1 Riverside Circle, Suite 102, Roanoke, Virginia 24016, (540)581-0121.

## Abstract

**Objectives:** Potentially harmful nonsteroidal anti-inflammatory drugs (NSAIDs) utilization persists at undesirable rates throughout the world. The purpose of this paper is to review the literature on interventions to de-implement potentially harmful NSAIDs in healthcare settings and to suggest directions for future research.

**Design:** Scoping review

**Data Sources:** PubMed, CINAHL, Embase, Cochrane Central, and Google Scholar (2000-2022)

**Study Selection:** Studies reporting on the effectiveness of interventions to systematically reduce potentially harmful NSAID utilization in healthcare settings.

**Data Extraction:** Using Covidence systematic review software, we extracted study and intervention characteristics, including the effectiveness of interventions in reducing NSAID utilization.

**Results:** From 7,818 articles initially identified, 68 were included in the review. Most studies took place in European countries (45.6%) or the U.S. (35.3%), with randomized controlled trial as the most common design (55.9%). The majority of studies (76.2%) reported a reduction in the utilization of potentially harmful NSAIDs. Interventions were largely clinician-facing (76.2%) and delivered in primary care (60.2%). Academic detailing, clinical decision support or electronic medical record interventions, performance reports, and pharmacist review were frequent approaches employed. NSAID use was most commonly classified as potentially harmful based on patients’ age (55.8%) or history of gastrointestinal disorders (47.1%) or kidney disease (38.2%). Only 7.4% of interventions focused on over-the-counter NSAIDs in addition to prescription. Few studies (5.9%) evaluated pain or quality of life following NSAIDs discontinuation.

**Conclusion:** Many varied interventions are effective in de-implementing potentially harmful NSAIDs in healthcare settings. Efforts to adapt, scale, and disseminate these interventions are needed. In addition, future interventions should address over-the-counter NSAIDs, which are broadly available and widely used. Evaluating unintended consequences of interventions, including patient-focused outcomes, is another important priority.

- **What is already known on this topic** – *NSAIDs are overutilized by high-risk patients at persistent rates. Interventions to reduce potentially harmful NSAIDs prescribing and over-the-counter (OTC) NSAIDs use are needed*.
- **What this study adds** – *More than 50 studies published internationally from 2000 to 2022 (three quarters of those reviewed) reported on interventions effective in de-implementing potentially harmful NSAIDs. We extracted and summarized characteristics of studies and of interventions to identify research gaps and priorities for future research*.
- **How this study might affect research, practice or policy** – *In light of the large number of effective interventions on record, implementation and dissemination efforts should be the priority of future work*.

## BACKGROUND

Nonsteroidal anti-inflammatory drugs (NSAIDs) reduce pain and inflammation through inhibition of cyclooxygenase (COX-1 and -2) enzymes, thereby limiting the production of inflammatory prostaglandins.^1^ Representing 5 to 10% of global medication utilization, NSAIDs are commonly used to treat arthritis and musculoskeletal pain, injuries, headache, and other sources of acute and chronic pain.^2^ There are six classes of NSAIDs: salicylates, propionic acid derivatives, acetic acid derivatives, enolic acid derivatives, anthranilic acid derivatives, and selective COX-2 inhibitors.^3^ NSAIDs are available in prescription and over-the-counter (OTC) strengths, a variety of different formulations, and oral (most common), intravenous, injectable, and topical forms. The use of NSAIDs has risen globally throughout the last twenty years,^4–8^ in part due to increasing rates of chronic and persistent pain and an increasing aging population.

In addition to anti-inflammatory and analgesic properties, NSAIDs have numerous other physiologic effects, which differ by NSAIDs class. For example, NSAIDs can reduce the integrity of the gastrointestinal mucosal barrier and limit submucosal blood flow, increasing risk of ulceration, hemorrhage, or perforation, particularly among vulnerable individuals; COX-2 selective NSAIDs are associated with lower gastrointestinal risk.^1,9^ Taking NSAIDs can reduce renal blood flow, alter fluid-electrolyte balance, and increase risk of acute kidney injury.^10^ Risk for and worsening of hypertension, heart failure, and other cardiovascular issues have also been associated with regular NSAIDs use.^10–13^ In 2015, the U.S. Food and Drug Administration updated the black box warning on OTC NSAIDS to include, “*NSAIDs can increase the risk of heart attack or stroke in patients with or without heart disease or risk factors for heart disease…The risk of heart attack or stroke can occur as early as the first weeks of using an NSAID…There is an increased risk of heart failure with NSAID use*.*”*^14^ Medical societies and professional organizations around the world have established recommendations for limiting or avoiding NSAIDs in certain high-risk populations **(Supplementary File 1)**.

Despite numerous long-standing recommendations, potentially harmful NSAIDs prescribing and OTC use persists globally.^15–18^ As an example, multiple studies show that up to 30% of patients with chronic kidney disease (CKD) are prescribed long-term NSAIDs.^19,20^ This high-risk use has resulted in a substantial number of adverse events; NSAIDs are a leading cause of drug-related hospitalizations and mortality.^21–23^ The drivers of potentially harmful NSAIDs prescribing and use are complex and multilevel.^24–26^ Clinicians’ unfamiliarity with professional recommendations, clinical inertia, limited alternative options for pain management, lack of patient knowledge or understanding, and broad availability of OTC NSAIDs are just some of the factors involved. The evolving regulatory landscape also complicates NSAIDs practice patterns and decision-making (a timeline of major NSAIDs-related regulatory events and other key historical timepoints in the U.S., as an example, is shown in **Table 1**).^27,28^

**Table 1.** Timeline of Major Regulatory Events and Other Key Historical Timepoints U.S. NSAID History.

| Year | Event |
| --- | --- |
| 1900 | Aspirin registered in the U.S., available via prescription.<sup>29</sup> |
| 1915 | Aspirin approved by FDA for over-the-counter distribution.<sup>30</sup> |
| 1964-1976 | Indomethacin, ibuprofen, diclofenac, ketoprofen, and naproxen approved by the FDA.<sup>31,32</sup> |
| 1971 | John Vane discovered the mechanism of action of aspirin and other NSAIDs.<sup>33</sup> |
| 1976 | COX enzyme discovered, recognized for role in prostaglandin synthesis.<sup>33</sup> |
| 1984 | Ibuprofen approved by FDA for over-the-counter distribution.<sup>34</sup> |
| 1985 | FDA approved aspirin for treatment of acute myocardial infarction and secondary cardiovascular prevention, CDC endorses.<sup>35</sup> |
| 1991 | Second COX enzyme ("COX-2") discovered, recognized as identical in structure but having important differences in substrate and inhibitor selectivity and in intracellular locations.<sup>36</sup> |
| 1997 | Celecoxib, the first selective COX-2 inhibitor, was introduced.<sup>37</sup> |
| 2004-2005 | Selective COX-2 inhibitors (rofecoxib and valdexocib) withdrawn from the market based on evidence that long-term use increases cardiovascular risk. Celecoxib remained on the market with a black box warning. The warning was also added to the over-the-counter NSAIDs' drug facts label.<sup>38</sup> |
| 2007 | FDA approved topical diclofenac at the prescription-level.<sup>39</sup> |
| 2015 | Strengthening of the black box warning over-the-counter NSAIDs' drug facts labels related to risk of heart attack and stroke.<sup>14</sup> |
| 2016 | The USPSTF recommends initiating low-dose aspirin use for the primary prevention of cardiovascular disease (CVD) and colorectal cancer (CRC) in adults aged 50 to 59 years (B recommendation).<sup>40</sup> |
| 2020 | Topical diclofenac approved for over-the-counter distribution.<sup>28</sup> |
| 2022 | Department of Health and Human Services initiates the Million Hearts Campaign, a national initiative to prevent 1 million heart attacks and strokes within 5 years. It focuses on implementing a set of evidence-based priorities that can improve cardiovascular health (including appropriate aspirin use).<sup>41</sup> |
| 2022 | The USPSTF recommends that for adults aged 40 to 50 years with an estimated 10% or greater 10-year cardiovascular disease (CVD) risk: The decision to initiate low-dose aspirin use for the primary prevention of CVD in this group should be an individual one. (C recommendation).<sup>42</sup> |

CDC: Centers for Disease Control and Prevention, COX: cyclooxygenase, CRC: colorectal cancer, CVD: cardiovascular disease, FDA: Food and Drug Administration, NSAIDs: nonsteroidal anti-inflammatory drugs, USPSTF: United States Preventive Services Task Force

Further efforts are needed to reduce the potential harm associated with prescription and OTC NSAIDs and promote safer pain management for high-risk patients. The purpose of this paper is to provide an overview of published interventions to de-implement potentially harmful NSAIDs in healthcare settings, to identify knowledge gaps, and to suggest opportunities for subsequent interventions and future research related to NSAIDs de-implementation.

## METHODS

We performed a scoping review of the scientific and gray literature reporting on interventions to de-implement NSAIDs in healthcare settings. Our review was guided by the PRISMA Extension for Scoping Reviews.^43^ As a scoping review, this review is not eligible for PROSPERO registration, but the protocol was posted at https://osf.io/ywe62/ in January 2022.

### Eligibility Criteria

Eligible studies were published in English between 2000 and 2022, employed any study design, and evaluated interventions administered with a goal of de-implementing potentially harmful NSAIDs in a healthcare setting.

**Healthcare settings** included any outpatient or inpatient healthcare environments within any medical specialty.

**NSAIDs** included prescription or over-the-counter oral or topical NSAIDs. NSAIDs that are not approved for current use were included if they had been approved at any point during the study period. For example, although rofecoxib and valdecoxib were removed from the U.S. market in 2005,^38^ they were included in the literature search. Aspirin taken for cardiovascular disease prophylaxis (<100 mg) was not included since the recommended dose is lower than that commonly used for analgesic purposes. If the purpose of the intervention was to study an inappropriate prophylactic use of aspirin, an exception was made to include that study. To be included, studies must have reported NSAID prescribing or use rates before and after the intervention, at minimum.

**Potentially harmful** NSAIDs included those that were prescribed or taken in a manner inconsistent with professional recommendations or otherwise recognized as high-risk by the study authors.

**Interventions** were defined as “any activity or set of activities aimed at modifying a process, course of action, or sequence of events in order to change one or several of their characteristics such as performance of expected outcome”, as described by the World Health Organization.^44^ Interventions were actively delivered to healthcare clinicians, healthcare teams, or directly to patients. All interventions included in the study involved de-implementation of NSAIDs. Passive interventions such as policy changes were not included.

**De-implementation** was defined as the systematic reduction or elimination of potentially harmful NSAID prescribing or use, or the modification of some aspect of NSAID prescribing or use to improve safety and/or reduce risk of harm (e.g., taking proton pump inhibitors in combination with NSAIDs).

**Patient populations** of focus were limited to adults >18 years of age and with any medical condition.

### Search Strategy

With the guidance of a professional librarian, we searched PubMed, CINAHL, Embase, Cochrane Central, Google Scholar and Google for [intervention OR program OR related MESH terms] + [de-implement OR deprescribe OR reduce OR related MESH term] + [nonsteroidal anti-inflammatory drug OR NSAID OR related MESH term] in Spring 2022 (full search strategy appears in **Supplementary File 2**). Studies were limited to articles or abstracts published between 2000 and 2022. Only studies written in or translated into English were included.

Studies identified in the search were uploaded as abstracts to Covidence (Melbourne, Australia), an online systematic review management platform. Duplicate studies were auto-identified by Covidence and deleted. Two members of the research team (MR and MA) independently screened all abstracts for inclusion in the review. Discrepancies were resolved by conference with a third team member (JE). Studies that passed the screening stage were moved to full text review. Three members of the research team (MR, MA, and ES) independently reviewed all full-text studies for alignment with eligibility criteria and reviewed reference lists for additional studies. Discrepancies were resolved via conference among the three reviewers.

Since the initial search identified some studies that focused on NSAIDs as one of multiple medications addressed in de-implementation or deprescribing interventions, we performed a second PubMed, CINAHL, Embase, Cochrane Central, and Google Scholar search for systematic and scoping review articles related to medication de-implementation or deprescribing published between 2000 and 2022. Abstracts identified in the search were reviewed by the lead author (MR) to eliminate reviews that did not meet inclusion criteria. Each review article was independently searched by two team members (EO and JT) for studies that included NSAIDs and met all other inclusion criteria. Discrepancies were resolved via conference among the two reviewers.

### Data Extraction

Studies that passed full-text review were moved to the charting/data extraction phase. Using the Covidence extraction framework, data were independently extracted by two team members (MA and ES). Two additional team members (MR and EO) downloaded the extracted data table from Covidence, independently checked a 25% data sample for accuracy, and resolved discrepancies by consensus.

The following data were extracted for each study: publication year, country, study design, intervention setting, type of intervention (de-implementation approach), intervention participants (e.g., physicians, pharmacists, patients), NSAIDs involved in intervention (prescription and/or OTC; classes and/or specific medications), and focus patient population. We also documented the effectiveness of the intervention in de-implementing NSAIDs (yes, no, or no change) and any patient-focused outcomes evaluated in relation to the intervention. Extracted data were integrated and synthesized into tables and figures based on data extraction elements listed above. The research team collectively appraised results to summarize the identified interventions and identify gaps in the literature.

## RESULTS

The original search identified 7,720 studies from which 60 were included in the final review. The secondary systematic and scoping review search identified 98 articles from which eight additional papers were included in the final review. **Figure 1** details the flow of articles through identification and screening stages and **Supplementary File 3** shows all articles included in the final review (n= 68).

**Figure 1.**
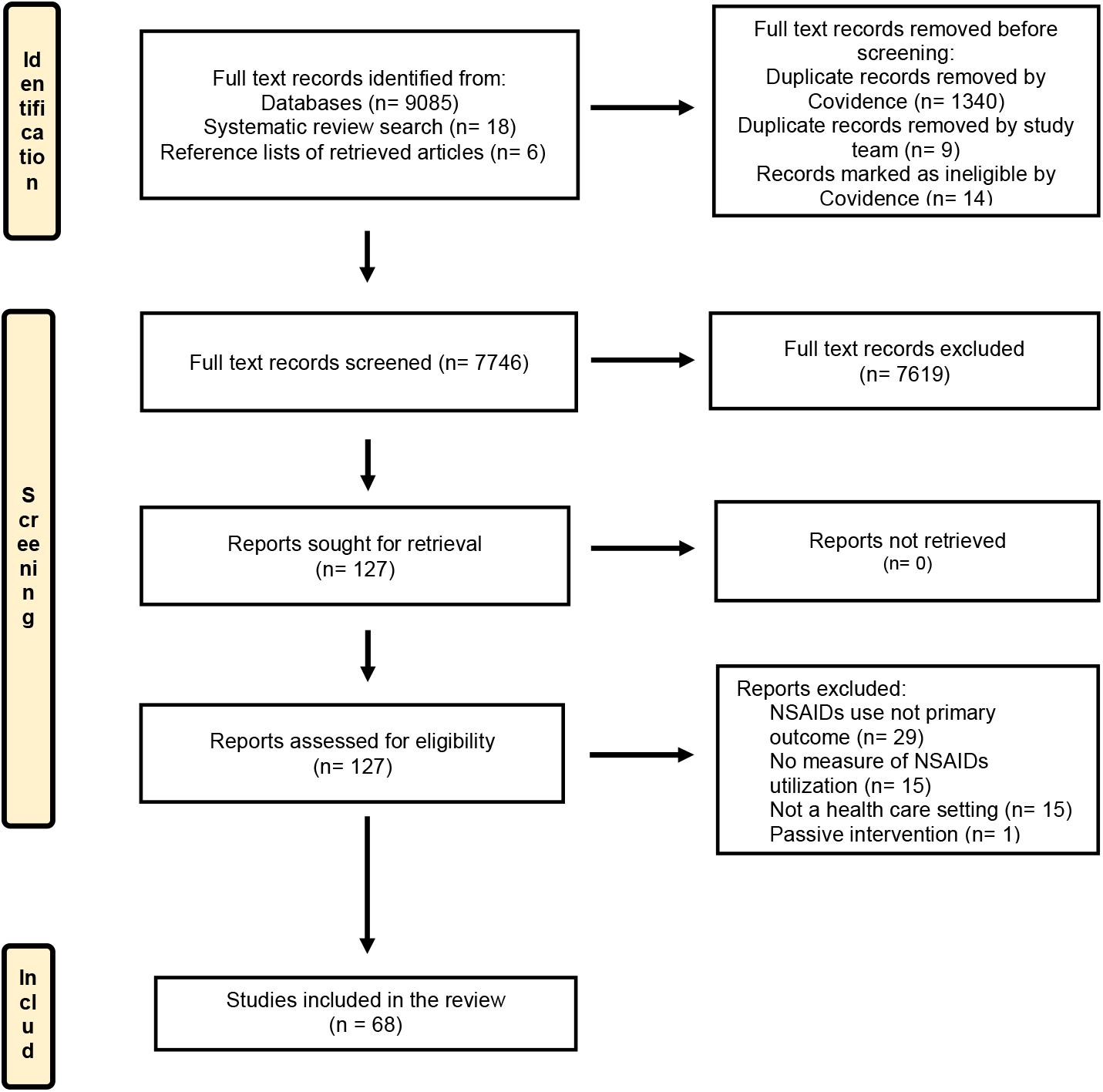
PRISMA Flow Diagram: Identification, Screening, and Included Studies in a Scoping Review of Interventions to De-implement Potentially Harmful NSAIDs in Health Care Settings.

### Characteristics of Studies

A total of 27 (39.7%) of studies were published between 2000 and 2010, with the remaining published between 2011 and 2022. The majority of studies took place in a European country (45.6%) or the United States (35.3%) (**Supplementary File 3**). A variety of study designs were represented, with randomized controlled trial (RCT) being the most common (55.9%) and prospective, interventional trials also frequently used (23.5%).

### Characteristics of Interventions

Most interventions were delivered to clinicians (i.e., clinician-facing) (76.5%) **(Supplementary File 3a)**, although some were patient-facing (8.8%) **(Supplementary File 3b)** and some were both clinician and patient-facing (10.3%) **(Supplementary File 3c)**. Of the clinician-facing and both clinician and patient-facing interventions, primary care or general practice physicians were the most frequent focus (72.6%), with pharmacists, nurses, and physicians in sub-specialty settings the focus of the remaining interventions. Both single component (54.4%) and multi-component (45.6%) interventions were employed. The most common intervention approach, represented in more than half of studies, was academic detailing and/or clinician education (**Figure 2**). Interventions focused on the electronic medical record (EMR) and/or clinical decision support were common among single component interventions, while clinician performance reports or audit/feedback and medication review by a pharmacist were common among multi-component interventions (**Figure 2**).

**Figure 2.**
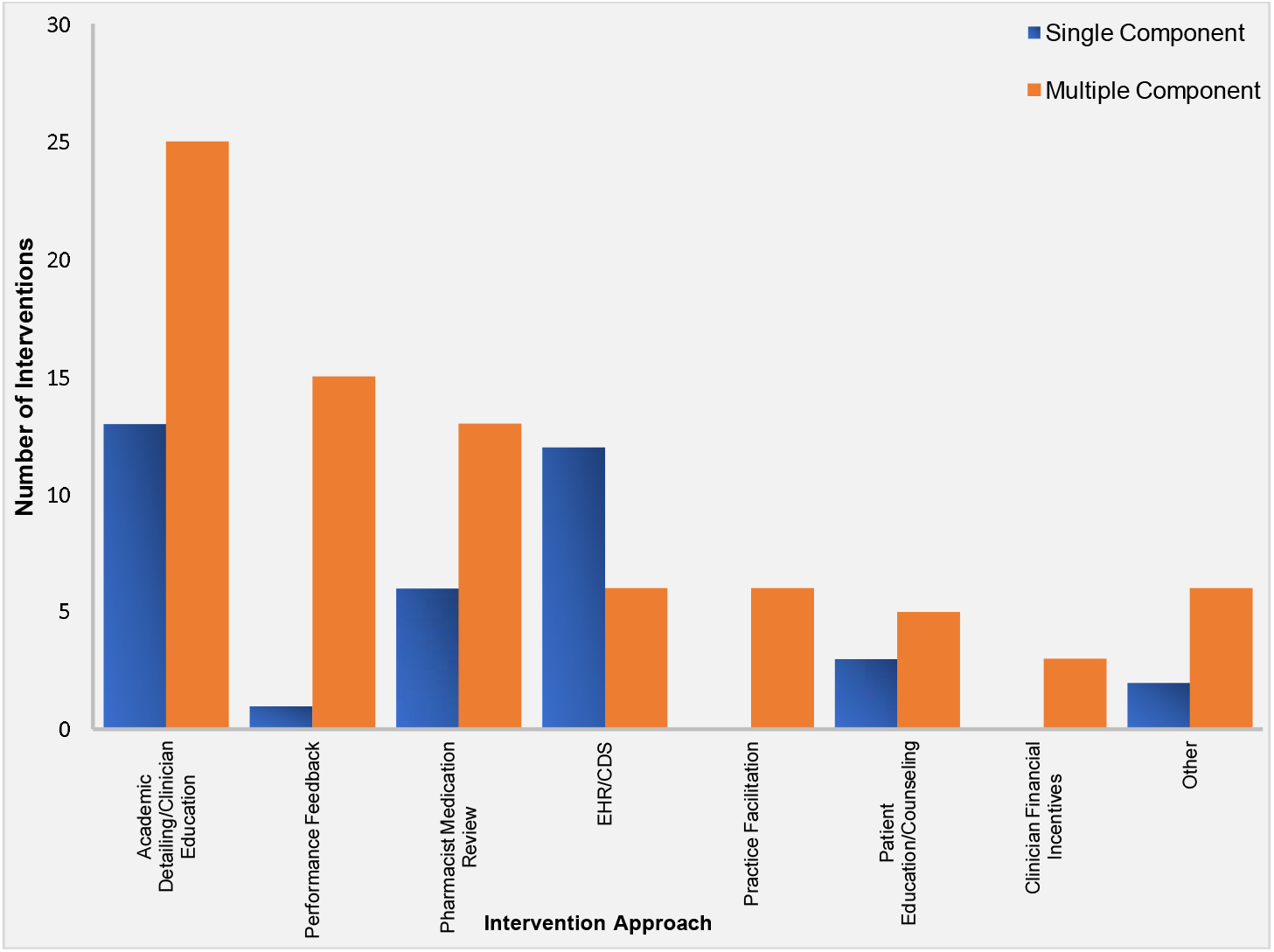
Single and Multiple Component Intervention Approaches to De-implement Potentially Harmful NSAIDs in Health Care Settings.

Some interventions focused solely on NSAIDs, while 26 (59.1%) focused on de-implementation of other medications as well. For example, the EQUiPPED trial^45^ aimed to reduce prescribing of multiple potentially harmful medications to older adults in the emergency department, while the study reported by Dreishulte et al.^46^ focused on de-implementation of high-risk NSAIDs and antiplatelet agents in primary care. Most interventions (85.2%) aimed to de-implement all types of NSAIDS, although some (14.8%) targeted reduction of a single type or class of NSAIDs. Interventions largely focused on prescription NSAIDs, with only 7.4% of interventions aimed to reduce potentially harmful OTC NSAIDs. All studies focused on oral NSAIDs; topical NSAIDs were not addressed in any interventions.

More than half of interventions (55.8%) aimed to de-implement NSAIDs classified as high-risk based on patient age (generally >65 or 70 years), with BEERS, START, and STOPP criteria frequently referenced (**Table 2**).^47^ The de-implementation of potentially harmful NSAIDs among patients with gastrointestinal conditions (e.g., peptic ulcer disease, inflammatory bowel disease) or who were taking chronic NSAIDs without gastroprotective medication (e.g., proton-pump inhibitor) and kidney disease was also common (47.1% and 38.2%, respectively) (**Table 2**).

**Table 2.** Criteria by which the Use of Nonsteroidal Anti-Inflammatory Drugs (NSAIDs) was Classified as High-Risk or Potentially Harmful by Included Studies.

| High-risk or potentially harmful due to: | Number of Studies (%) |
| --- | --- |
| Age (generally $>65$ or 70 years) | 38 (55.9) |
| Existing gastrointestinal conditions or lack of gastroprotective medication | 32 (47.1) |
| Kidney disease | 26 (38.2) |
| Cardiovascular disease or heart failure | 22 (32.3) |
| Hypertension | 19 (27.9) |
| Potential for medication interaction | 16 (23.5) |
| Lower risk alternative available | 10 (14.7) |
| Contribution to polypharmacy | 7.0 (10.3) |
| Chronic or long-term use | 6.0 (8.8) |
| Use of multiple medications containing NSAIDs | 4.0 (5.9) |

NSAIDs: nonsteroidal ant-inflammatory drugs

Most interventions (76.2%) were effective in reducing use of high-risk NSAIDs **(Supplementary File 3)**. Very few studies (5.9%) evaluated patients’ level of pain or quality of life following discontinuation of NSAIDs. Over half of studies (51.5%) assessed other patient-focused outcomes associated with the interventions, including patient-rated quality of interaction with clinician,^48^ occurrence of falls,^49^ and emergency department admissions.^46^

## DISCUSSION

Although many professional organizations and societies recommend limiting or avoiding the use of NSAIDs in high-risk patients, potentially harmful prescribing and OTC use persists at undesirable rates.^15–18^ This scoping review identified more than 50 studies (approximately three quarters of those reviewed) published between 2000 and 2022 that describe healthcare-based interventions effective in de-implementing potentially harmful NSAIDs. Future research should prioritize methods to adapt, scale, and disseminate these effective interventions. Such efforts stand to improve medication safety for older adults and patients with chronic conditions and other risk factors.

The interventions we reviewed spanned more than 20 years. During this time, there was a great deal of evolution in the NSAID market, in the scientific evidence related to the comparative effectiveness and safety of various NSAIDs and other analgesics, and in professional recommendations, clinical practice patterns, and regulatory policy related to NSAIDs prescribing. As such, iterative and rapid adaptations to the context, setting, and resources available are crucial to preserve internal validity and ensure the effectiveness of interventions. In addition to informed adaptation and implementation approaches, the science of de-implementation continues to evolve.^50–52^ Theories, frameworks, and models for de-implementation can complement and enhance the effective interventions we reviewed, the majority of which did not report being informed by any sort of implementation or de-implementation model.

Since the interventions described in this review varied widely in terms of de-implementation approaches employed, high-risk conditions addressed, populations of interests, and NSAIDs of focus, it is difficult to directly compare studies. However, it is notable that both single and multi-component interventions were both effective in de-implementing NSAIDs, which is inconsistent with the literature for many other low-value services.^53^ In several cases, interventions involving low-cost, low-burden approaches (e.g., one-time education session, online training modules, pamphlets) were associated with the same reduction in NSAIDs utilization as much more elaborate and costly approaches (e.g., pharmacist medication review, individual patient counseling, EMR workflow modification). A better understanding of intervention approaches and factors associated with effective de-implementation in different settings and for various high-risk scenarios would provide valuable insight to future efforts to improve NSAIDs safety.

We observed four additional important gaps in the literature related to NSAIDs de-implementation in healthcare settings. First, patient-facing interventions were infrequently employed. Direct engagement with patients can enhance outcomes of deprescribing and other health services interventions,^54–56^ and may be especially germane to the de-implementation of OTC NSAIDs, which were barely addressed by interventions studied. Second, outcomes important to patients were inconsistently assessed in the studies reviewed. Despite some evidence that patient satisfaction and trust are not adversely impacted by low-value care de-implementation,^57,58^ clinicians continue to cite concern about patient response as a predominant de-implementation barrier.^24,59–61^ In addition to evaluating patient-focused outcomes, future studies should explore unintended consequences of the interventions. Of the minority of studies that evaluated adverse events or changes in pain following NSAIDs de-implementation, none showed increases in adverse event or pain outcomes.^62–66^ In fact, one study reported lower pain levels among older adults who reduced NSAIDs as part of a pharmacist review program.^62^ Finally, although the reviewed interventions varied in duration, sustainability of the observed effects beyond the conclusion of active interventions was infrequently evaluated.

This study has some limitations. Our initial search strategy did not consistently identify studies that focused on general healthcare deprescribing or interventions to reduce multiple medications. To incorporate these studies into our review, we added a supplemental secondary database search which was effective in identifying additional applicable studies. Additionally, our data extraction plan did not capture whether interventions focused on reducing new prescriptions for (or OTC use of) potentially harmful NSAIDs vs. reducing refills for ongoing inappropriate NSAIDs, which could be important to informing future interventions. Last, as a scoping review, we did not formally evaluate the quality of the studies reviewed.

## CONCLUSION

This scoping review identified 68 interventions to de-implement potentially harmful NSAIDs published internationally from 2000 to 2022. Over three-quarters of the interventions were effective in reducing NSAIDs utilization. These interventions classified NSAID use/prescribing as high-risk for multiple reasons, employed a variety of de-implementation approaches, and took place in several different healthcare settings. Patient-facing interventions were under-represented and only two interventions included OTC NSAIDs. Few studies evaluated the sustainability of intervention outcomes or unintended consequences of interventions. Considering the large number and diversity of effective interventions on record, future efforts should prioritize the adaptation, scaling, and dissemination initiatives to de-implement high-risk NSAIDs and enhance medication safety for older adults and patients with chronic conditions.

## Data Availability

All data produced in the present study are available upon reasonable request to the authors.

## Footnotes

## Acknowledgements

The authors thank C. Cozette Comer, MS (University Libraries, Virginia Tech) for assistance with designing the literature search and Meera Abrishami and Laura Miller for assisting with the initial review of abstracts.

## Author Contributions

MSR and JWE designed the study; MSR and CCC designed the search strategy; MSR, JWE, and EGO designed the data extraction approach; MSR, EGO, JKT, and ES completed abstract review and data extraction; IY and MV developed manuscript tables; MSR wrote the first draft of the manuscript; all authors edited and revised the manuscript and approved the final version.

## Patient and public involvement

Patients were not involved in this research.

## Provenance and peer review

This article has not been previously published or peer reviewed.

