## Supplementary Files 1-3 for "A Scoping Review of Interventions to De-implement Potentially Harmful Nonsteroidal Anti-inflammatory Drugs (NSAIDs) in Healthcare Settings"

**Supplementary File 1. Sample Recommendations and Prescribing Notes from Professional Medical Societies and Organizations Related to Potentially Harmful NSAIDS**

| Organization | Recommendation Year | Recommendation |
| --- | --- | --- |
| American Association of Family Physicians (AAFP).^1^ | 2009 | - When possible, NSAIDs should be avoided in persons with preexisting renal disease, congestive heart failure, or cirrhosis. - Consider monitoring serum creatinine levels after initiation of NSAID therapy in persons at risk of renal failure, and in those taking angiotensin-converting enzyme inhibitors and angiotensin receptor blockers. - NSAIDs and aspirin should be avoided in persons taking anticoagulants. If concurrent NSAID and anticoagulant use is necessary, an increase in INR should be anticipated. There should be appropriate INR monitoring and warfarin (Coumadin) dosage adjustments, and GI prophylaxis should be initiated. - Asthma could be induced or worsened as a result of taking NSAIDs. - Ibuprofen, indomethacin, and naproxen (Naprosyn) are safe to use in breastfeeding women. |
| American Geriatric Society Beer’s Criteria.^2^ | 2023 | In older adults:   - Avoid chronic use of NSAIDs unless other alternatives are not effective and patient can take gastroprotective agent (proton pump inhibitor or misoprostol). - Avoid short-term scheduled use in combination with corticosteroids, anticoagulants, or antiplatelet agents unless other alternatives are not effective and the patient can take a gastroprotective agent. - Use with caution in patients with heart failure who are asymptomatic; avoid in patients with symptomatic heart failure: Dronedarone NSAIDs and COX-2 inhibitors. - In patients with kidney disease and Cr/Cl <30ml/min, avoid NSAIDs (non-selective, COX-2 selective, and nonacetylated salicylates, oral and parenteral) may increase the risk of acute kidney injury and a further decline in kidney function |
| American Heart Association (AHA).^3^ | 2007 | - NSAIDs should be taken at lowest effective dosage for the shortest duration possible to reduce cardiovascular risk. - COX inhibitors carry the highest cardiovascular risk and thus, naproxen is recommended as the drug of choice for patients with cardiovascular risk. |
| American Society of Nephrology (ASN)/ Choosing Wisely.^4^ | 2012 | - Avoid NSAIDs in individuals with hypertension or heart failure or CKD of all causes, including diabetes. |
| Arthritis Society of Canada.^5^ | 2022 | - Do not use NSAIDs before, during or after heart surgery (bypass surgery). - Patients with a history of cardiovascular disease should be careful using NSAIDs. - Patients with risk factors for cardiovascular disease (e.g., diabetes, smoking, elevated cholesterol, obesity and family history) should also be careful using NSAIDs. Safer alternative treatments should be used if available. - NSAIDs should be used in the lowest effective dose for the shortest possible duration of time. |
| Chinese Pharmaceutical Association Hospital Pharmacy Professional Committee, Asia-Pacific Experts on Topical Analgesics Advisory Board.^6,7^ | 2018, 2022 | - Best available evidence indicates that topical NSAIDs have a moderate effect on relief of osteoarthritic pain, comparable to that of oral NSAIDs but with a better risk-to-benefit ratio. International clinical practice guidelines recommend topical NSAIDs on par with or ahead of oral NSAIDs for pain management in patients with osteoarthritis, and as the first-line choice in persons aged ≥75 years. |
| European Alliance of Associations for Rheumatology (EULAR).^8^ | 2021 | - NSAIDs, at the lowest effective dose, should be added or substituted in patients who respond inadequately to paracetamol. In patients with increased gastrointestinal risk, non-selective NSAIDs plus a gastroprotective agent, or a selective COX-2 inhibitor, should be used. |
| Health Canada.^9^ | 2021 | - Advises pregnant women to not use NSAIDs from 20 to 28 weeks of pregnancy, unless advised by a health care professional, due to risk of kidney damage and low amniotic fluid. - NSAIDs are contraindicated for use during the third trimester of pregnancy because of risk of premature closure of the ductus arteriosus and the potential to prolong parturition. |
| Kidney Disease Improving Global Outcomes (KDIGO).^10^ | 2012 | - Avoid NSAIDs in people with GFR <30 ml/min/1.73 m^2^. - Prolonged NSAID therapy is not recommended in people with GFR <60 ml/min/1.73 m^2^. - NSAIDs should not be used in people taking lithium. - Avoid NSAIDs in people taking RAAS blocking agents. |
| Medicines and Health care Projects Regulatory Agency (MHRA) (UK). ^11,12^ | 2009, 2015 | - Patients at risk of renal impairment or renal failure (particularly elderly people) should avoid NSAIDs if possible - if NSAID treatment is absolutely necessary, then the lowest effective dose for the shortest possible duration should be used to control symptoms - the renal function of such patients should be carefully monitored during NSAID treatment. - It is important to consider other concomitant disease states, conditions, or medicines that may precipitate reduced renal function when prescribing NSAIDs |
| National Institute for Health and Care Excellence (NICE).^13^ | 2013 | - NSAIDs should be prescribed with caution as courses of just a few days, even at doses within prescribing recommendations, can be associated with serious adverse effects in susceptible patients. - In primary care, paracetamol is recommended in preference to NSAIDs, where appropriate. If a patient is likely to benefit from NSAID treatment naproxen or ibuprofen are recommended first-line, at the lowest effective dose, for the shortest possible time. Patients taking NSAIDs who are at increased risk of complications require regular monitoring. |
| NHS Clinical guideline.^14^ | 2019 | - Avoid NSAIDs in in severe cardiac failure, hepatic failure, and active peptic ulcer disease. - Concomitant use of NSAIDs and other nephrotoxics (e.g., ACE Inhibitors, Angiotensin Receptor Blockers, lithium, and diuretics) should be avoided where possible to prevent the risk of acute kidney injury. - Use caution with NSAIDs in the elderly and with hepatic insufficiency and mild renal impairment. - Avoid combinations of NSAIDs. - Alcohol consumption and cigarette smoking are possible lifestyle risk factors for serious NSAID-induced gastrointestinal adverse effects. |
| North American Spine Society (NASS).^15^ | 2020 | - Non-selective NSAIDs are suggested for the treatment of low back pain. - There is insufficient evidence to make a recommendation for or against the use of selective NSAIDs for the treatment of low back pain. |
| Society of Hospital Pharmacists of Australia (SHPA).^16^ | 2018 | - NSAIDs should be avoided before any surgery where postoperative bleeding would be of concern. - COX-2 selective NSAIDs may be used preoperatively as they have limited effect on platelet function. |
| STOPP/START Criteria.^17^ | 2023 | The following prescriptions are potentially inappropriate to use in patients aged 65 years and older:   - Long-term systemic i.e., non-topical NSAIDs with known history of coronary, cerebral or peripheral vascular disease (increased risk of thrombosis). - NSAIDs or systemic corticosteroids with heart failure requiring loop diuretic therapy (risk of exacerbation of heart failure). - Long-term aspirin at doses greater than 100mg per day (increased risk of bleeding, no evidence for increased efficacy). - NSAIDs and vitamin K antagonist, direct thrombin inhibitor or factor Xa inhibitors in combination (risk of major gastrointestinal bleeding). - NSAIDs if eGFR < 50 ml/min/1.73m2 (risk of deterioration in renal function). - NSAIDs other than COX-2 selective agents with history of peptic ulcer disease or gastrointestinal bleeding, unless with concurrent PPI or H2 antagonist (risk of peptic ulcer relapse). - NSAIDs with severe hypertension i.e., systolic blood pressure consistently above 170 mmHg and/or diastolic blood pressure consistently above 100 mmHg (risk of exacerbation of hypertension). - Long-term use of NSAID (>3 months) for symptom relief of osteoarthritis pain where paracetamol has not been tried (simple analgesics preferable and usually as effective for pain relief and safer). - Long-term NSAID or colchicine (>3 months) for chronic treatment of gout where there is no contraindication to a xanthine-oxidase inhibitor. - NSAID with concurrent corticosteroids for treatment of arthritis/rheumatism of any kind (increased risk of peptic ulcer disease). |

ACE: angiotensin-converting enzyme, CKD: chronic kidney disease, COX: cyclooxygenase, eGFR/GFR: estimated glomerular filtration rate/glomerular filtration rate, H2: histamine 2-receptor, GI: gastrointestinal, INR: international normalization ratio, NSAIDs: nonsteroidal ant-inflammatory drugs, PPI: proton pump inhibitors, PRN: pro re nata, RAAS: renin–angiotensin–aldosterone system.

**Supplementary File 2. Terms Used in Literature Search**

| **NONSTEROIDAL ANTI-INFLAMMATORY DRUGS** | **DE-IMPLEMENTATION** | **INTERVENTION** |
| --- | --- | --- |
| Acetylsalicylic acid  Advil  Aleve  Anti-inflammatory  Anti-inflammatory agent  Aspirin  Bextra  Celebrex  Celocoxib  Daypro  Diclofenac  Etodolac  Etoricoxib  Fenoprofen  Flurbiprofen  Ibuprofen  Indocin  Indomethacin  Ketoprofen  Ketorolac  Lodine  Mefanamic acid  Meloxicam  Motrin  Nabumetone  Nalfon  Naproxen  Non-opiate  Non-opioid  Nonsteroidal anti-inflammatory drugs  Nonsteroidal anti-inflammatory medication  NSAID  Oxaprozin  Piroxicam  Ponstel  Relafen  Rovecoxib  Salicylic acid  Steroidal, non  Sulindac  Toradol  Valdecoxib  Vioxx  Voltaren | Cease, ceasing, ceased  Decrease, decreasing. decreased  De-escalate, de-escalating, decreased, de-escalation  De-implement, de-implementing, de-implemented, de-implementation  Deimplement, deimplementing, deimplementation, deimplementation  Deprescribe, deprescribing  Discontinue, discontinuing, discontinued, discontinuation  Mitigate, mitigating, mitigated, mitigation  Phase out, phasing out, phased out  Reduce, reducing, reduced, reduction  Remove, removing, removed, removal  Stop, stopping, stopped  Taper, taper off, tapering, tapered  Terminate, terminating, terminated, termination  Withdraw, withdrawing, withdrawn, withdrawal | Academic detailing  Audit and feedback  Clinical decision support  CME  Continuing medical education  Counseling  Education  Inappropriate prescribing  Initiative  Intervention  Measure  Medication review  Pharmacist counseling  Pharmacist review  Program  Strategy  Electronic medical record tool  Electronic medical record strategy  EMR tool  EMR strategy  Electronic health record tool  Electronic health record strategy  EHR tool  EHR strategy  Financial incentive  Performance feedback  Performance improvement  Performance incentive  Performance report  Practice coaching  Practice facilitation  Quality  Quality improvement  Safety |

**Supplementary File 3. Articles Included in the Final Review (n=68)**

| **Supplementary File 3a. Clinician Facing Interventions (n=55)** | | | | | |
| --- | --- | --- | --- | --- | --- |
| **Study title** | **Author**  **(Year)** | **Country** | **Health care setting** | **Type of intervention** | **Intervention reduced NSAIDs use?** |
| - A pharmacist-led information technology intervention for medication errors (PINCER): a multicenter, cluster-randomized, controlled trial and cost-effectiveness analysis.^18^ | Avery  (2012) | United Kingdom | General Practice/ Primary Care | Pharmacist-led information technology intervention composed of clinician education, feedback, and dedicated support | Yes |
| - CURATA: A patient health management program for the treatment of osteoarthritis in Québec: An integrated approach to improving the appropriate utilization of anti-inflammatory/analgesic medications.^19^ | Beaulieu  (2004) | Canada | General Practice/ Primary Care | Clinician education workshop | Yes |
| - Evidence-based educational outreach visits: Effects on prescriptions of non-steroidal anti-inflammatory drugs.^20^ | Bernal-Delgardo (2002) | Spain | General Practice/ Primary Care | Academic detailing | Yes |
| - Improving ambulatory prescribing safety with a handheld decision support system: A randomized controlled trial.^21^ | Berner  (2006) | United States | General Practice/ Primary Care | A personal digital assistant (PDA)–based clinical decision support system | Yes |
| - Influencing NSAID prescribing in primary care using different feedback strategies.^22^ | Braybrook (2000) | United Kingdom | General Practice/ Primary Care | Clinician education, active or passive practice-specific prescribing feedback, and prescribing workbook | Yes |
| - The implementation of academic detailing and its effectiveness on appropriate prescribing of pain relief medication: A real-world cluster randomized trial in Belgian General Practice/ Primary Care.^23^ | Bruyndonck  (2018) | Belgium | General Practice/ Primary Care | Academic detailing | Yes |
| - A group randomized trial to improve safe use of nonsteroidal anti-inflammatory drugs.^24^ | Curtis  (2005) | United States | Managed Care Organization | Continuing medical education and audit & feedback with peer-derived benchmarks | No |
| - Safer prescribing — A trial of education, informatics, and financial incentives.^25^ | Dreischulte (2016) | Scotland | General Practice/ Primary Care | Clinician education, computerized clinical decision support, and financial incentives to practices to review patients’ charts for appropriateness of NSAIDs use | Yes |
| - Academic detailing as a method of continuing medical education.^26^ | Dyrkorn  (2019) | Norway | General Practice/ Primary Care | Academic detailing | Yes |
| - A large-scale initiative to improve NSAID prescribing safety.^27^ | Eskildsen  (2017) | New Zealand | General Practice/ Primary Care | Practice facilitation, including academic detailing, workflow coaching, performance feedback, and other safe prescribing resources | Yes |
| - Computerized clinical decision support during medication ordering for long-term care residents with renal insufficiency.^28^ | Field  (2009) | Canada | Long-term Care Facility | Computerized clinical decision support system alerts | Yes |
| - One-to-one versus group sessions to improve prescription in primary care: A pragmatic randomized controlled trial.^29^ | Figueiras  (2001) | Spain | General Practice/ Primary Care | One-on-one and group clinician education and reminders | Yes |
| - Prevention of potentially inappropriate prescribing for elderly patients: A randomized controlled trial using STOPP/START criteria.^30^ | Gallagher  (2011) | Ireland | Inpatient Care | Screening by pharmacist using STOPP/START criteria and follow-up visit with primary care | Yes |
| - Impact of EHR-based clinical decision support on adherence to guidelines for patients on NSAIDs: A randomized controlled trial.^31^ | Gill  (2011) | United States | General Practice/ Primary Care | EHR-based clinical decision support alerts for high-risk patients | Yes |
| - Effect of an academic detailing intervention on the utilization rate of cyclooxygenase-2 inhibitors in the elderly.^32^ | Graham  (2008) | Canada | Tertiary Medical Center | Academic detailing | Yes |
| - Guided medication dosing for elderly emergency patients using real-time, computerized decision support.^33^ | Griffey  (2012) | United States | Tertiary Medical Center | Computerized decision support tool | Yes |
| - Data feedback and behavioral change intervention to improve primary care prescribing safety (EFIPPS): Multicenter, three-arm, cluster randomized controlled trial.^34^ | Guthrie  (2016) | Scotland | General Practice/ Primary Care | Emailed educational material with support for identifying high-risk patients or feedback on high-risk prescribing, with or without a behavioral change component | Yes |
| - A physician-focused intervention to reduce potentially inappropriate medication prescribing in older people.^35^ | Keith  (2013) | Italy | General Practice/ Primary Care | Academic detailing, alternative drug list for potentially avoidable medications, prescribing reviews | Yes |
| - Reducing inappropriate non-steroidal anti-inflammatory prescription in primary care patients with chronic kidney disease.^36^ | Keohane  (2017) | Ireland | General Practice/ Primary Care | Automated EHR alert | Yes |
| - Toward safer prescribing: Evaluation of a prospective drug utilization review system on inappropriate prescriptions, prescribing patterns, and adverse drug events and related health expenditure in South Korea.^37^ | Kim  (2018) | South Korea | General Practice/Primary   Care | Drug utilization review system | No |
| - The prevalence of ’triple whammy’ prescriptions in surgical inpatients and associated pharmacist recommendations.^38^ | Koeck  (2021) | Germany | Inpatient Surgical Wards | Pharmacist medication review | Yes |
| - Pharmacist-led medication review in patients over 65: A randomized, controlled trial in primary care.^39^ | Krska  (2001) | Scotland | General Practice/ Primary Care | Pharmacist medication review | Yes |
| - Interdisciplinary geriatric and psychiatric care reduces potentially inappropriate prescribing in the hospital: Interventional study in 150 acutely Ill elderly patients with mental and somatic comorbid conditions.^40^ | Lang  (2012) | Switzerland | Inpatient Medical-Psychiatric Unit | Integrated care (a daily collaboration between a geriatrician and a psychiatrist providing interdisciplinary health  care management) | No |
| - Effectiveness of an academic detailing intervention in Primary Care on the prescribing of non-steroidal anti-inflammatory drugs.^41^ | Langaas  (2019) | Norway | General Practice/ Primary Care | Academic detailing | Yes |
| - Effects of an intervention (SÄKLÄK) on prescription of potentially inappropriate medication in elderly patients.^42^ | Lenander  (2017) | Sweden | General Practice/ Primary Care | Clinician self-assessment, peer review & feedback, and written change agreements | Yes |
| - Evaluation of a complex intervention to improve primary care prescribing: A phase IV segmented regression interrupted time series analysis.^43^ | MacBride-Stewart  (2017) | Scotland | General Practice/ Primary Care | Clinician education, performance feedback, pharmacist support, and  financial incentives | Yes |
| - Improving medication use in newly admitted home health care patients: A randomized controlled trial.^44^ | Meredith  (2002) | United States | Home Health Care | Medication improvement program (pharmacist consultations with home health nurse) | No |
| - Leveraging practice-based research networks to accelerate implementation and diffusion of chronic kidney disease guidelines in primary care practices: A prospective cohort study.^45^ | Mold  (2014) | United States | General Practice/ Primary Care | Practice facilitation, including academic detailing and performance feedback | Yes |
| - Improving quality of NSAID prescribing by internal medicine trainees with an educational intervention.^46^ | Naughton  (2010) | United States | Ambulatory Care Internal Medicine | Clinician education program, local practice data & consensus conferences, polypharmacy journals, and audit & feedback | Yes |
| - Can a practice pharmacist improve prescribing safety and reduce costs in polypharmacy patients? A pilot study of an intervention in an Irish general practice setting.^47^ | Ó Ciardha  (2022) | Ireland | General Practice/ Primary Care | Pharmacist conducting holistic medication reviews in the study group over a 6-month period. | Yes |
| - Effectiveness of educational outreach visits compared with usual guideline dissemination to improve family physician prescribing—an 18-month open cluster-randomized trial.^48^ | Pinto  (2018) | Portugal | General Practice/ Primary Care | Clinician education outreach visits or online resources | No |
| - A quality use of medicines program for general practitioners and older people: A cluster randomized controlled trial.^49^ | Pit  (2007) | Australia | General Practice/ Primary Care | Academic detailing, medication risk assessment, performance feedback, and financial incentives | Yes |
| - Education to reduce potentially harmful medication use among residents of assisted living facilities: A randomized controlled trial.^50^ | Pitkala  (2014) | Finland | Assisted Living Facilities | Nurse education and training | Yes |
| - Reporting of estimated glomerular filtration rate: Effect on physician recognition of chronic kidney disease and prescribing practices for elderly hospitalized patients.^51^ | Quartarolo  (2007) | United States | Inpatient Care | GFR reporting | No |
| - Randomized trial to improve prescribing safety in ambulatory elderly patients.^52^ | Raebel  (2007) | United States | Health Maintenance Organization. | Medication prescribing alerts | No |
| - Impact of a general practitioner educational intervention on osteoarthritis treatment in an elderly population.^53^ | Rahme  (2005) | Canada | General Practice/ Primary Care | Clinician education workshop and prescribing decision tree | Yes |
| - Clinically important drug-drug interactions in poly-treated elderly outpatients: A campaign to improve appropriateness in general practice.^54^ | Raschi  (2015) | Italy | General Practice/ Primary Care | Academic detailing | Yes |
| - Educational program for physicians to reduce use of non-steroidal anti-inflammatory drugs among community-dwelling elderly persons: A randomized controlled trial.^55^ | Ray  (2001) | United States | Community Health Care | Clinician education program | Yes |
| - Outcomes of a randomized controlled trial of a clinical pharmacy intervention in 52 nursing homes.^56^ | Roberts  (2001) | Australia | Long-term Care Facility | Clinical pharmacy interventions in nursing homes - (relationship building, nurse education, and medication review) | Yes |
| - Potentially inappropriate prescribing to older patients: Criteria, prevalence, and an intervention to reduce It: The prescription peer academic detailing (Rx-PAD) study – A cluster-randomized, educational intervention in Norwegian general practice.^57^ | Rognstad  (2018) | Norway | General Practice/ Primary Care | Academic detailing | Yes |
| - A multifactorial intervention to lower potentially inappropriate medication use in older adults in Argentina.^58^ | Schapira  (2021) | Argentina | General Practice/Primary Care | Clinician education workshops, deprescribing algorithms, and email alerts | Yes |
| - Computerized prescribing alerts and group academic detailing to reduce the use of potentially inappropriate medications in older people.^59^ | Simon  (2006) | United States | General Practice/ Primary Care | Academic detailing and computerized alerts | No |
| - Educational program for nursing home physicians and staff to reduce use of non-steroidal anti-inflammatory drugs among nursing home residents: A randomized controlled trial.^60^ | Stein  (2001) | United States | Long-term Care Facility | Clinician education | Yes |
| - Enhancing Quality of Provider Practices for Older Adults in the Emergency Department (EQUiPPED).^61^ | Stevens  (2017) | United States | Emergency Department | Clinician education, clinical decision support, individual performance feedback | Yes |
| - Randomized clinical trial of a customized electronic alert requiring an affirmative response compared to a control group receiving a commercial passive CPOE alert: NSAID—warfarin co-prescribing as a test case.^62^ | Strom  (2010) | United States | Inpatient Care | EHR alerts | No |
| - The medical office of the 21st Century (MOXXI): Effectiveness of computerized decision-making support in reducing inappropriate prescribing in primary care.^63^ | Tamblyn  (2003) | Canada | General Practice/ Primary Care | Computerized clinical decision support | No |
| - Effectiveness of interventions by community pharmacists to reduce risk of gastrointestinal side effects in nonselective nonsteroidal anti-inflammatory drug users.^64^ | Teichert  (2014) | Netherlands | Pharmacies | Performance feedback | Yes |
| - Computerized decision support to reduce potentially inappropriate prescribing to older emergency department patients: A randomized, controlled trial.^65^ | Terrell  (2009) | United States | Emergency Department | Computerized clinical decision support | Yes |
| - Intervention to improve appropriate prescribing and reduce polypharmacy in elderly patients admitted to an internal medicine unit.^66^ | Urfer  (2016) | Switzerland | Inpatient Care/Internal Medicine Unit | Clinical decision support checklist tool | Yes |
| - A cluster randomized trial to measure the impact on nonsteroidal anti-inflammatory drug and proton pump inhibitor prescribing in Italy of distributing cost-free paracetamol to osteoarthritic patients.^67^ | Vicentini  (2019) | Italy | General Practice/ Primary Care | Clinician education | No |
| - Electronic health record alerts decreased non-steroidal anti-inflammatory drug prescriptions in patients with congestive heart failure: A quality improvement initiative.^68^ | Vincent  (2020) | United States | Inpatient Care | EHR alerts | Yes |
| - Guidelines and educational outreach visits from community pharmacists to improve prescribing in general practice: A randomized controlled trial.^69^ | Watson  (2001) | England | General Practice/ Primary Care | Clinician education plus mailed printed guidelines | No |
| - Assessment of clinical pharmacy interventions to reduce outpatient use of high-risk medications in the elderly.^70^ | Weddle  (2017) | United States | General Practice/ Primary Care | Pharmacist medication review and electronic alerts | Yes |
| - Estimated GFR reporting is associated with decreased nonsteroidal anti-inflammatory drug prescribing and increased renal function.^71^ | Wei  (2013) | Scotland | General Practice/ Primary Care | GFR reporting | Yes |
| - Pharmacist-led provider education on inappropriate NSAID prescribing rates.^72^ | Whitner  (2020) | United States | General Practice/ Primary Care | Academic detailing | Yes |
| **Supplementary File 3b. Patient Facing Interventions (n=6)** | | | | | |
| - Pharmacist counseling and the use of nonsteroidal anti-inflammatory drugs by older adults.^73^ | Bear  (2017) | United States | General Practice/ Primary Care | Counseling from pharmacist | Yes |
| - A nurse‐delivered advice intervention can reduce chronic non‐steroidal anti‐inflammatory drug use in general practice: A randomized controlled trial.^74^ | Jones  (2002) | United Kingdom | General Practice/ Primary Care | Advice and education from nurse | Yes |
| - Can a nurse-directed intervention reduce the exposure of patients with knee osteoarthritis to nonsteroidal anti-inflammatory drugs?^75^ | Mazzuca  (2004) | United States | Health Maintenance Organization | Advice and education from nurse about non-pharmacological self-management of osteoarthritis | Yes |
| - Evaluating the effectiveness of a patient storytelling DVD intervention to encourage patient-physician communication about nonsteroidal anti-inflammatory drug (NSAID) use.^76^ | Miller  (2016) | United States | General Practice/ Primary Care | Video modeling patient-physician communication about NSAID use | No |
| - Effect of mobile device-assisted N-of-1 trial participation on analgesic prescribing for chronic pain: Randomized controlled trial.^77^ | Odineal  (2020) | United States | General Practice/ Primary Care | Mobile educational app | Yes |
| - Evaluation of a pharmacist-managed nonsteroidal anti-inflammatory drugs deprescribing program in an integrated health care system.^78^ | Rashid  (2020) | United States | Integrated Health System | A pharmacist managed NSAID deprescribing program | Yes |
| **Supplementary File 3c. Clinician and Patient-Facing Interventions (n=7)** | | | | | |
| - Randomized controlled trial of an intervention to improve drug appropriateness in community-dwelling poly-medicated elderly people.^79^ | Campins  (2017) | Spain | General Practice/ Primary Care | Medication evaluation program | Yes |
| - Effectiveness of a multifaceted intervention for potentially inappropriate prescribing in older patients in primary care: A cluster-randomized controlled trial (OPTI-SCRIPT Study).^80^ | Clyne  (2015) | Ireland | General Practice/ Primary Care | Academic detailing, pharmacist medicine review, and tailored patient information leaflets | No |
| - Pharmacist intervention reduces gastropathy risk in patients using NSAIDs.^81^ | Ibanez-Cuevas (2008) | Spain | Pharmacies | Structured interviews with patients and performance feedback reports with clinicians | Yes |
| - Effect of a pharmacist-led educational intervention on inappropriate medication prescriptions in older adults: The D-PRESCRIBE randomized clinical trial.^82^ | Martin  (2018) | Canada | Pharmacies | Pharmacist-generated educational brochure to patients and deprescribing recommendation to physicians | Yes |
| - Effect of pharmaceutical care services on outcomes for home care patients with heart failure.^83^ | Triller  (2007) | United States | Hospital/ Home Health care | Pharmacist in-home medication review with patients and feedback/recommendations to clinicians | No |
| - Safer Prescribing and Care for the Elderly (SPACE): A pilot study in general practice.^84^ | Wallis  (2018) | New Zealand | General Practice/ Primary Care | Academic detailing, audit & feedback, and educational mailings to patients | Yes |
| - NSAID use after bariatric surgery: A randomized controlled intervention study.^85^ | Yska  (2016) | Netherlands | Inpatient Surgical Health Care | Mailings to patients and their general practitioners on  the risks of NSAIDs after bariatric surgery | No |

CPOE: commercial computerized provider order entry, EHR: electronic health record, GFR: glomerular filtration rate, NSAIDs: nonsteroidal ant-inflammatory drugs, Rx-PAD: prescription peer academic detailing, STOPP/START: screening tool of older persons’ potentially inappropriate prescriptions/Screening tool to alert to right treatment.

13.	National Institute for Health and Care Excellence (NICE). Non-steroidal anti-inflammatory drugs. Manchester: NICE; 2013. Available from: www.nice.org.uk (Accessed Sep, 2013).

14.	NHS guidelines NSAIDs. Accessed July 12, 2023. https://www.nhsggc.org.uk/media/254733/oral-non-steroidal-anti-inflammatory-nsaid-guideline.pdf

15.	Ridge B. Diagnosis & Treatment of Low Back Pain.

68.	Vincent LT, Jacobs M, olarte neal, et al. Abstract 255: Electronic Health Record Alerts Decreased Non-Steroidal Anti-Inflammatory Drug Prescriptions in Patients With Congestive Heart Failure: A Quality Improvement Initiative. *Circ Cardiovasc Qual Outcomes*. 13(Suppl_1):A255-A255. doi:10.1161/hcq.13.suppl_1.255
